# Whether core stability training has a positive therapeutic effect on LBP patients: a Meta-analysis

**DOI:** 10.1101/2023.05.22.23290316

**Authors:** Zecheng Li, Xuebin Liu, Siya Li

## Abstract

**Objective:** The main goal of this study is to determine whether engaging in core stability exercises benefits LBP sufferers.

**Background:** As a common exercise therapy, core stability training has gradually become the main treatment for LBP patients to relieve pain. Although many similar researches have showed a significant impact of core stability training on LBP, many scholars still have the opposite experimental conclusion, that is, core stability training has no significant therapeutic effect on LBP patients.

**Methods:** Only randomized controlled could be included in this study, and we used the Web of Science, Cochrane Library, Embase, CNKI databases, China Science and Technology Journal Database and PubMed for article retrieval. Among them, non-LBP patients, non-core training, and articles with imperfect outcome indicators were not included.

**Results:** This analysis incorporated findings from 21 relevant studies in total. The results showed that, although that the overall effect was not significant, core stability training was helpful for LBP patients. The results of two secondary outcomes (generic health and specific function) showed that core stability training had no practical significance for the improvement of generic health and specific function of LBP patients. The specific results are as follows: pain (SMD = 2.74, 95%CI: 1.40-4.08, P < 0.0001), disability (SMD = 2.52, 95%CI: 1.69-3.36, P <0.00001), generic health (SMD = 1.08, 95%CI: 0.07-2.08, P = 0.04), specific function (SMD = 1.99, 95%CI: −0.04-4.02, P = 0.05).

**Conclusions:** We recommend that core stability training be used for short-term therapy, but not for long-term therapy.

## Introduction

Low back pain (LBP) is a frequent illness that impacts people’s daily activities and work. It can be roughly divided into specific LBP (SLBP), non-specific LBP (NSLBP or NLBP), acute LBP, chronic LBP (CLBP), chronic non-specific LBP (CNLBP) and recurrent LBP. Some scholars have proposed in related studies that LBP has a serious and far-reaching impact on people, so it is difficult to assess its impact on people’s daily lives[1]. Frist, LBP will affect 80% of people in their lifetime, and the lifetime prevalence rate is over 80%. Secondly, when receiving LBP treatment, it will affect the daily work of patients, increase the risk of other diseases and additional medical expenses while causing wage losses, to a certain extent, damage the social labor force and endanger social development [2, 3].

According to the research of Xu [4], they divided the main clinical treatment methods of LBP into drug therapy, physical factor therapy, and exercise therapy. Because drug therapy may be accompanied by side effects such as nausea and fatigue [5], physical factor therapy is difficult to treat long-term pain, so exercise therapy will gradually become the mainstream treatment of LBP. After summarizing past research and clinical practice, other scholars also found that exercise therapy appears to be one of the effective means to reduce pain, promote recovery in LBP patients [6, 7]. Based on this, Dianne Liddle conducted a questionnaire survey on exercise therapy for LBP patients in Ireland. The questionnaire includes many types of exercise techniques, such as spinal muscle-related exercises, conventional muscle exercises, and specialized, customized exercises [8], to establish exercise therapy recommendations and exercise rehabilitation guidelines for chronic LBP management.

There are many kinds of exercise therapy for LBP, such as routine exercise, ball stability exercise, muscle strengthening exercise, Pilates exercise and so on. Among them, core stability exercise is not only a very popular and easy to practice fitness method, but also an important treatment measure in the field of rehabilitation medicine and sports medicine [9]. Despite the fact that several similar research have demonstrated a significant impact of core stability training on LBP [10, 11]. However, in recent years, many scholars have obtained the opposite conclusion in clinical trials. After comparing core stability training with regular training, Shamsi argued that traditional training is just as beneficial as core stability training for reducing disability and pain in LBP patients, and there is no significant difference in the therapeutic effect between traditional training and core stability training [12]. Similar to Shamsi, Smith also concluded that core stability training had no significant effect on LBP patients after including 29 related studies up to 2013 [2]. In summary, different scholars have different attitudes towards the therapeutic effect of core stability training, and the influence of core stability exercise on LBP patients is controversial.

Shojania [13] was proposed in 2007 that about 23% of review studies will expire within two years. Therefore, different scholars ’ different views and opinions on the therapeutic effect of core stability training may be caused by timeliness. In addition, previous studies were different in the selection of subjects and the setting of intervention measures. For example, Wang’s restrictive definition of core stability exercises is to practice on unstable surfaces, in contrast to other scholars who chose core muscle workouts and core-related stable motions, and the quantity of literatures included is small, with only 5 papers included [3]; Niederer and Mueller[14] [14] only evaluated individuals with chronic nonspecific LBP and found no benefit of core stability training on other categories of LBP patients. According to this, we conclude that one of the reasons why scholars have different views on the therapeutic effect of core stability training is that the types of interventions included in the study and control group are too single.

Based on the above problems, this study retrieved and included the latest RCT results in recent years to ensure the timeliness of the study. At the same time, a variety of LBP patients including NSLBP, CLBP, and recurrent LBP were widely included in the study to prevent the simplification of the study subjects from affecting the results. In terms of intervention measures, we adopt a broader definition of the experimental group, including core stability training, core strength training, core muscle strengthening training, segmental stability and other exercises related to core stability, and in the control group also included various exercise treatments. The purpose is to explore whether core stability exercise therapy is superior to other exercise therapies, and whether core stability exercise has a good therapeutic effect and positive impact on all kinds of LBP patients.

## methods

This review was structured using the PRISMA method (Preferred Reporting Items for Meta-Analyses and Systematic Reviews). The procedure was also included in the International Prospective Registry of Systematic Reviews. (PEOSPERO: CRD42023404448).

### search strategy

We used the following Boolean search syntax to search for potential related articles, the complete search strategy sees Table 1. We deleted duplicate documents retrieved in multiple databases.

Table 1. search strategy

### Inclusion Criteria

Each study had to meet the PICO standards in order to be taken into consideration for this review.

#### Types of studies

Just the randomized controlled trial (RCT) was examined in this study. Studies comparing a collection of stabilization exercises with various physical therapy or medical therapy groups were also included. We only included the research with Chinese and English as a language.

#### Types of participants

We excluded studies that included participants with Lumbar surgery. Also, we did not include any patients whose LBP was brought on by particular diseases or disorders. Meanwhile, we did not set any limitations for age and gender.

#### Types of interventions and comparisons

As mentioned above, we only accept core stability or core stability-related training as the intervention group. We have no restrictions on the control group, including various exercise therapies, drug therapies, and many different physical therapies.

Only one control group’s data can be reviewed if the study includes two or more control groups (for instance, general exercise and massage treatment or general exercise and massage therapy and muscle strengthening exercise).

#### Types of outcome measures

The primary outcomes of this review were pain intensity and disability, and the visual analog scale was predominantly used to generate the pain intensity measure (VAS). The outcome unit we accepted is 1-100mm. If the unit is not uniform, we will perform unit conversion when analyzing the data.

For disability, most studies which we included used the Oswestry Disability Questionnaire (ODQ), the Modified Oswestry Disability Questionnaire (MODQ) and the Oswestry Disability Index (ODI) to assess disability. And only a small part of studies used other tools to evaluate disability, such as the Roland Morris Disability Questionnaire (RMDQ), and the Functional Rating Index (FRI) questionnaire.

The secondary outcomes were patient-specific function and generic health. We evaluated generic health by using Short-Form 36 (SF-36) and measured function by using Patient-Specific Functional Scale (PSFS).

### Study selection and data extraction

Using predetermined criteria, we chose the titles, abstracts, and full papers of pertinent studies. Then we extracted the following data from the included articles: study design, information of participants (age, gender, numbers), intervention design (experimental and control), treatment condition (frequency and duration), outcome measure, follow-up, and drop-out. Afterwards, we created a common table by using these data (S2 Table.).

Table 2. Characteristics of Included Studies

### Assessing the Risk of Bias

We evaluated the risk of bias for each article using the Cochrane Handbook for Systematic Reviews of Interventions [15]. To assess the caliber of the included research, we also employed the PEDro scale, which has 11 components [16]. If two reviewers dispute the results of the study, consult a third reviewer for a decision.

### Statistical Analysis

Using Review Manager Software, we conducted analyzes on each of the papers we included (RevMan5.2). Because the data included in the study are continuous data, we pooled the data, chose the standardized mean difference (SMD) as a useful sign, and provided a 95% confidence interval (CI) for the variance.

The I^2^ statistic was used to quantify heterogeneity, and the Cochrane Q statistic was used to determine whether heterogeneity occurred among the included studies (test level = 0.05). Since our data is random, we chose the random-effects (RE) model. If the level of heterogeneity is too high, we will do subgroup analysis and sensitivity analysis to identify the factors contributing to it. If it is impossible to pinpoint the exact cause of heterogeneity, a descriptive analysis is conducted.

## Result

### Search Results

As previously noted, we search studies in the Embase, Cochrane Library, PubMed, Web of Science, and CNKI databases. Fig 1 depicts the entire selection procedure for qualifying studies.

Fig 1. Research and study selection for PRISMA-compliant systematic reviews.

We initially retrieved 964 related articles from the database. 632 articles were excluded before screening, of which 413 were duplicates, 77 were excluded by automated tools, and 142 were excluded for other reasons. After reviewing the title, abstract, and keywords of relevant studies, reviewers rejected 302 papers. and discovered that there were 6 reports that couldn’t be retrieved. After reading the full text of the included studies, we excluded 3 articles, reasons being: Incomplete data[17, 18](n = 2), Ineligible controls [19](n = 1). That left a total of 21 studies for inclusion[12, 17, 20-39].

### Characteristics of the included studies

For the primary outcomes, as shown in Supplementary Table 2, 21 studies of LPB patients were eligible, with 16 studies used a visual analogue scale (VAS) to measure pain. 4 studies used the ODI to measure disability, whilst 4 studies measured disability by using the RMDQ, 2 studies used the ODQ to measure disability, and two studies measured disability by using the MODQ. One study also included the FRI questionnaire as a disability outcome measure.

For the secondary outcomes, we included 2 studies for measuring specific function by using the Patient-Specific Functional Scale (PSFS). And we still included 2 studies as a generic health outcome measure, which used the Short-Form 36 (SF-36).

### Risk of bias and quality assessment

The PEDro scale revealed that none of the studies were of low quality (see S4 Table.) and Fig 2 depicts the risk of bias for the included research. The PEDro scale showed that although only 5 articles used the blind method (blinding of participants or therapists or assessors) in the research process, the overall score was still high, and most participants were randomly assigned and hidden during allocation.

Table 3. Study quality and risk of bias.

Fig 2. Risk of bias of this review

One study of the 21 studies included in this review did not use randomized techniques and did not report allocation concealment[28]. In addition, we also found one study did not report complete outcome data[36]. We believed that the design features of the two studies may affect the results of the experiment. Hence, it was determined that the two studies had a significant probability of bias.

### Outcomes

#### pain intensity

17 studies used VAS to assess the efficacy of core stability training in LBP patients. Through the forest plot generated by related software, the VAS scale was significantly higher in the experimental group than in the control group (SMD = 2.98, 95%CI: 1.79-4.18, P < 0.00001). Then we performed a subgroup analysis of the included studies using duration as a criterion due to the high heterogeneity. It was divided into 5 subgroups and we excluded 5 studies by sensitivity analysis[25, 30, 34, 36, 39].

The results show that the heterogeneity between some groups has decreased slightly, but the overall heterogeneity is still high. The subgroup heterogeneity of 8 weeks duration (I^2^ = 44%) decreased, the heterogeneity of the 6-week subgroup did not change significantly (I^2^ = 96%), as well as a shift in the total pooled effect (SMD = 2.74, 95%CI: 1.40-4.08, P < 0.0001). See Fig 3 for all details.

Fig 3. forest plot of pain intensity

#### disability

We performed subgroup analysis based on different outcome measures tool (Fig 4). In addition, we performed a sensitivity analysis because the findings of the study were highly heterogeneous, and we ultimately decided to exclude 4 related studies[22, 25, 26, 35]. The results of ODI (SMD = 3.66, 95%CI: 2.76-4.55, P < 0.00001), RMDQ (SMD = 1.66, 95%CI: 0.54-2.79, P = 0.004) and FRI (SMD = 2.68, 95% CI: 1.85-3.50, P < 0.00001) showed that core stability exercise significantly improved LBP patients, but ODQ (SMD = 3.22, 95%CI: −0.64-7.08, P = 0.10) and MODQ (SMD = 2.83, 95%CI: −1.12-6.79, P = 0.16) were not statistically significant.

Fig 4. forest plot of disability

#### generic health

We calculated the physical component score and mental component score of patients by using the SF-36, and the results indicated that there is a significant degree of heterogeneity in the outcome indicators. As Fig 5 shown that both physical component score (SMD = 1.64, 95%CI: −1.75-5.04 P = 0.34) and mental component score (SMD = 0.73, 95%CI: −0.04-1.51, P = 0.06) were not statistically significant.

Fig 5. forest plot of generic health

#### specific function

Fig 6 shows that there were only 2 studies included in this review for specific function, result have high heterogeneity (SMD = 1.99, 95%CI: −0.04-4.02, I^2^ = 98%) and not statistically significant (P = 0.05). There was no subgroup analysis of the results because there weren’t many literatures included.

Fig 6. forest plot of specific function

## discussion

### Summary of main findings

The results of pain intensity (SMD = 2.74, 95% CI: 1.40-4.08), disability (SMD = 2.68, 95%CI: 1.85-3.50), health (SMD = 1.08, 95%CI: 0.07-2.08) and function (SMD = 1.99, 95%CI: −0.04-4.02) were all shown that core stability training has a beneficial impact and effect on those with LBP. This view is consistent with the results of a meta-analysis published by Smith, Wang, Han [2, 3, 6, 14, 40-42]. In terms of follow-up, not all studies have followed up on the patients, but the analysis of the known follow-up results shows that there are no adverse reactions to the treatment of LBP patients with core stable exercise.

### Sustainable therapeutic effect

This review does not examine the effect of core stability exercises on the long-term sustainability of LBP patients, as Coulombe and Elbayomy did. Only the results of the pain intensity part involve the sustainable therapeutic effect of core stability training on LBP patients. Summing up the existing results, the experimental group’s VAS data results with interventions lasting six, eight, and twelve weeks were considerably better than those with interventions lasting four and three months. This shows that the short- and medium-term advantages of core stability training outweigh the long-term advantages, which is consistent with the previous research results[2,3,40,41].

### Therapeutic effect of all LBP patients

Due to the lack of relevant studies included in this review, subgroup analysis cannot be performed on all types of LBP patients. On the whole, the types of LBP patients included in the review are rich, involving NLBP, CNLBP, subacute NSLBP, acute NSLPB, recurrent LBP. According to the findings, core stability training has different effects on different types of LBP patients. The impact on some LBP patients may not be significant, but on the whole, it can still have a positive impact on LBP patients, which is basically consistent with the meta-analysis results of other single-type LBP patients. In summary, we believed that core stability training has a positive effect on all types of LBP patients, and compared with other treatment methods, core stability training also has a certain improvement effect in reducing the pain and disability impact of LBP patients.

### Explanation of heterogeneity

The heterogeneity of the four outcome indicators in this review is high, so we performed sensitivity analysis and subgroup analysis on the two primary outcome indicators (pain and disability). Although the heterogeneity between subgroups is slightly reduced, the heterogeneity of the overall results is still high (pian intensity: I^2^ = 97%, disability: I^2^ = 95%). The intervention strategies used in the control group and the diversity of participants were the main differences between the studies included in this study, as shown in Table 2. Considering the goal of this study, we cannot optimize it, so we believe that the above two points are important reasons for the high heterogeneity of the main outcome indicators of this study.

The heterogeneity of the results of the two secondary outcome indicators is also too high. We believe that the reasons for the high heterogeneity of the secondary outcome indicators should be like the reasons for the high heterogeneity of the main outcome indicators. In addition, insufficient relevant studies to allow us to perform sensitivity analysis and subgroup analysis may also be an important factor leading to high heterogeneity.

### Limitation of this review

There were several limitations to this review that should be mentioned. The overall number of studies included in this review was too few to do a sufficient subgroup analysis, which is the first limitation. The second limitation is the high heterogeneity. Due to the purpose of the study, the participants and interventions of the experiment cannot be unified, so the high heterogeneity may affect the credibility of the final results.

## Conclusion

We feel that core stability training is effective for treating low back pain and has a good impact on patients with all types of LBP based on the examination of the research findings. However, it is only suitable for reducing the pain of LBP patients and reducing the impact of disability on daily life. The findings of this study cannot conclusively demonstrate that core stability training also benefits LBP patients in other ways (general health, particular function). Compared with other exercise therapies, we believe that core stability training has advantages in the treatment of LBP patients, but the advantages are not obvious. For short-term therapy, but not for long-term therapy, we advise using core stability exercises.

## Author contributions

Zecheng Li: main writer of this review, work for study design, data collection, data extraction, data analysis and draft preparation.

Xuebin Liu: work for study design, data collection.

Siya Li: work for study design, data extraction.

## Data Availability

Data openly available in a public repository.

## Supporting information

**S1 Table.** search strategy.

| Search term |  |
| --- | --- |
| #1 | core exercise or core training or core stability exercise* or core stability training or core stability or “core stability exercise therapy” or core stability strength training or core stabilization training |
| #2 | nonspecific low back pain* or Chronic Low Back Pain* or LBP or low back pain or NLBP or CNLBP or NSLBP |
| #3 | health or physical fitness* or health status* or fitness, medical* or healthy or healthiness or well-being* |
| #4 | #1 AND #2 AND #3 |

**S2 Table.**
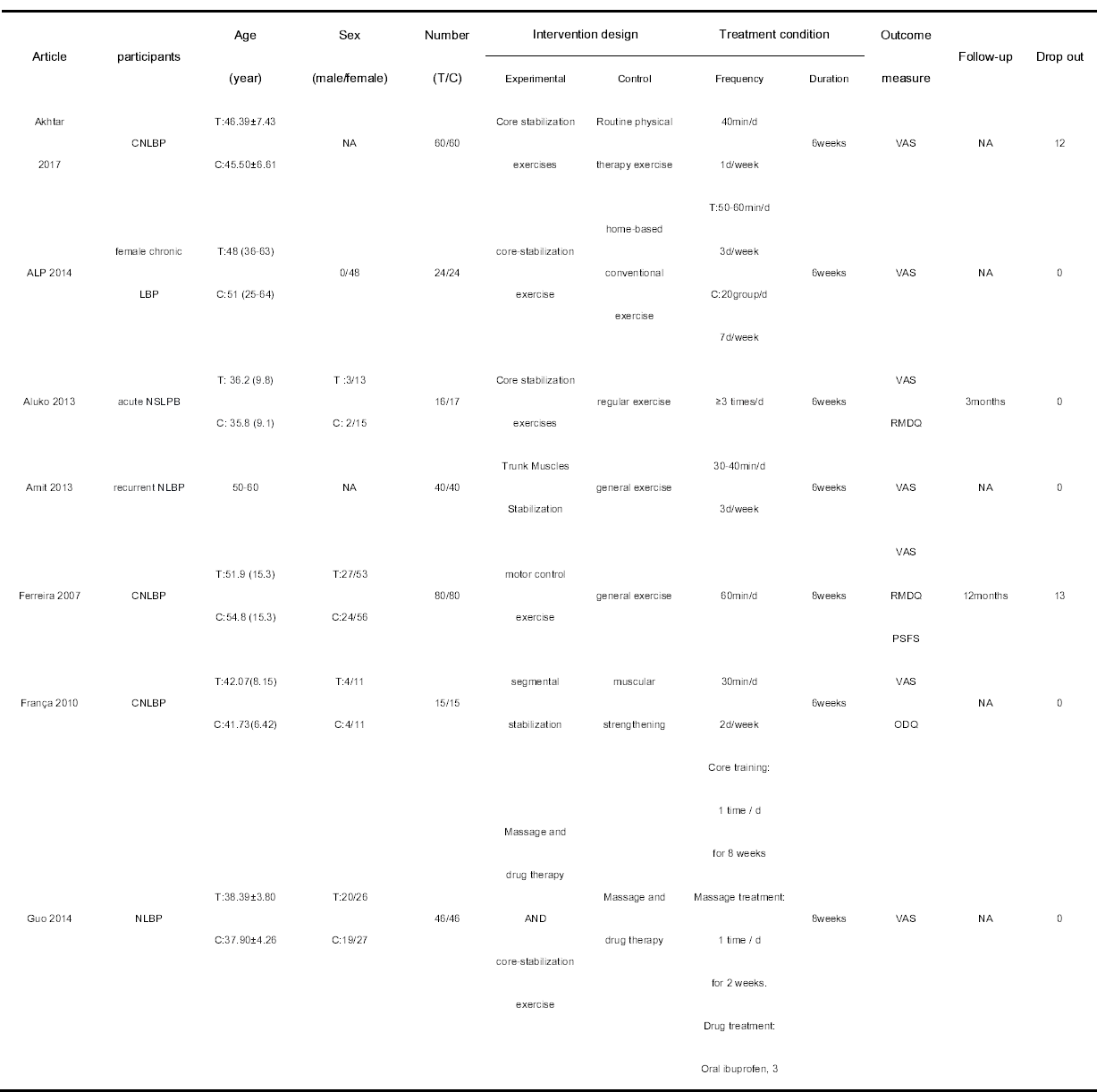
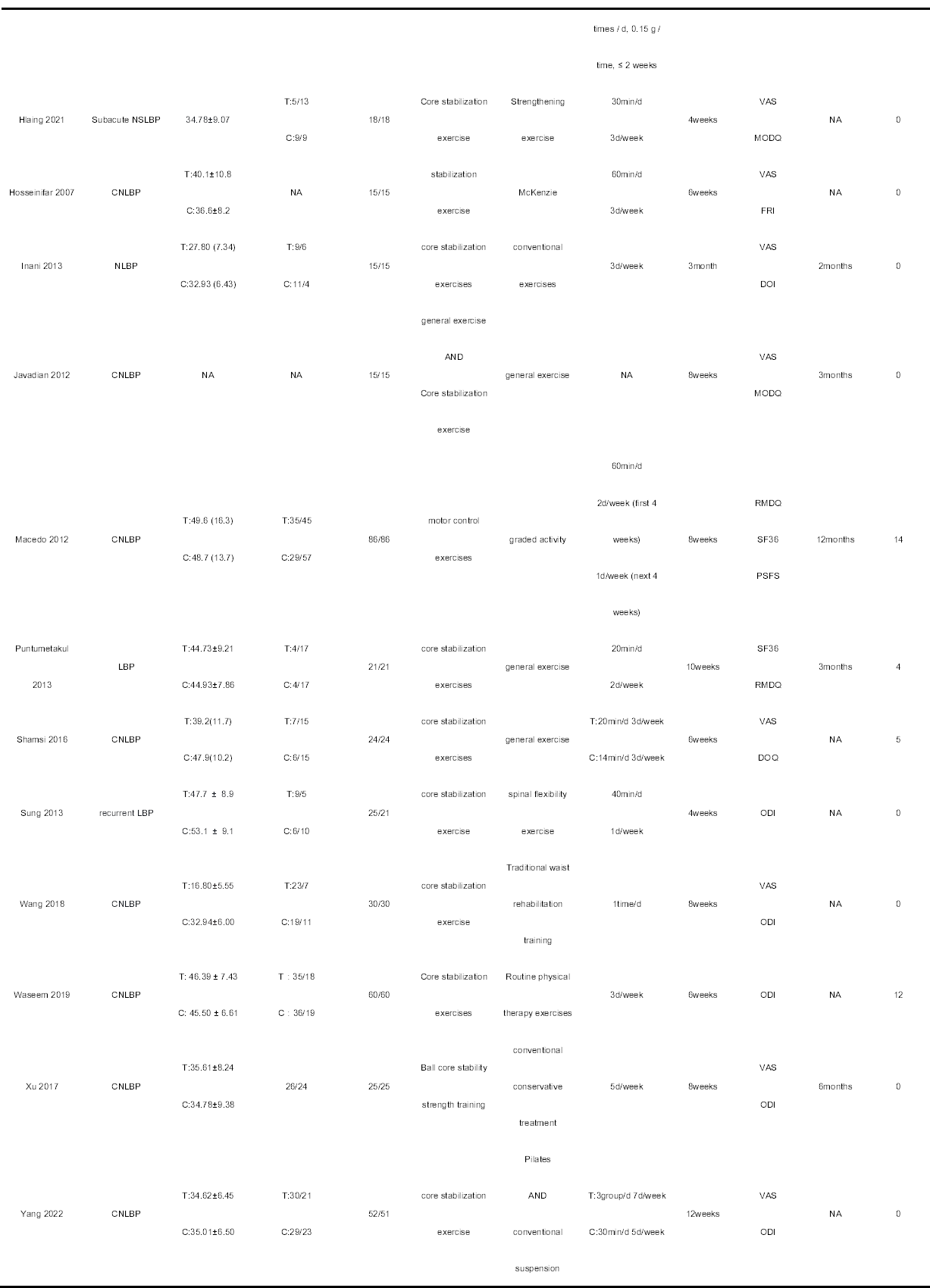
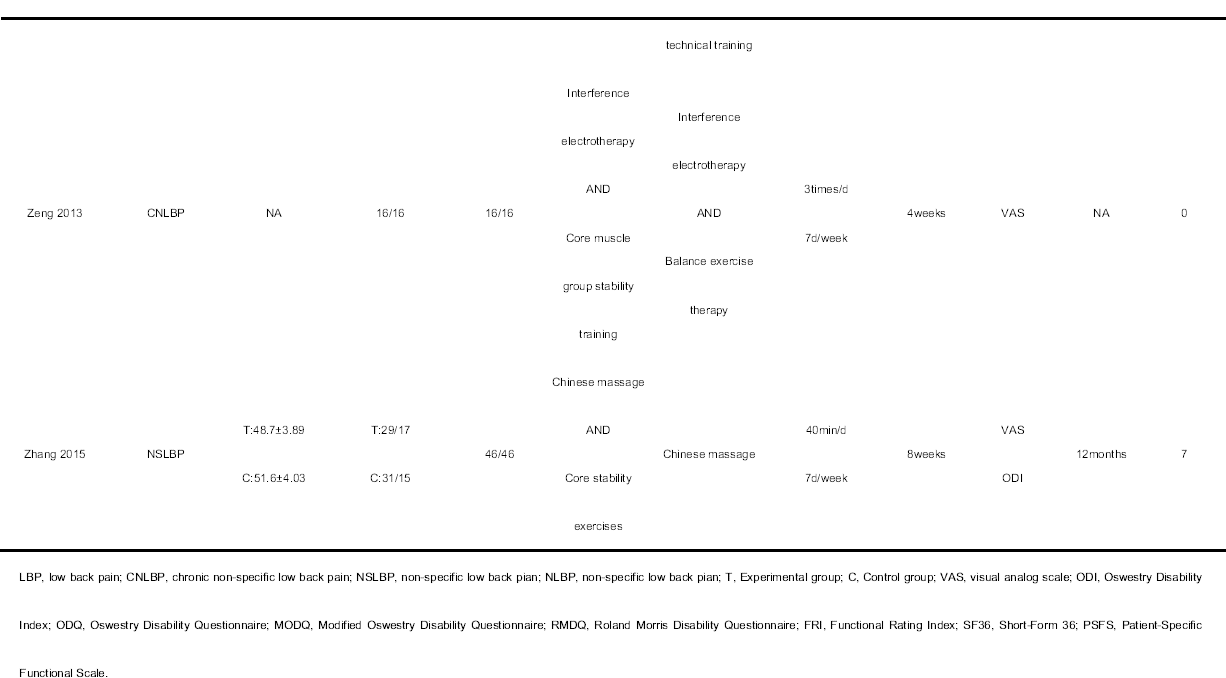
Characteristics of Included Studies.

**S3 Figure.**
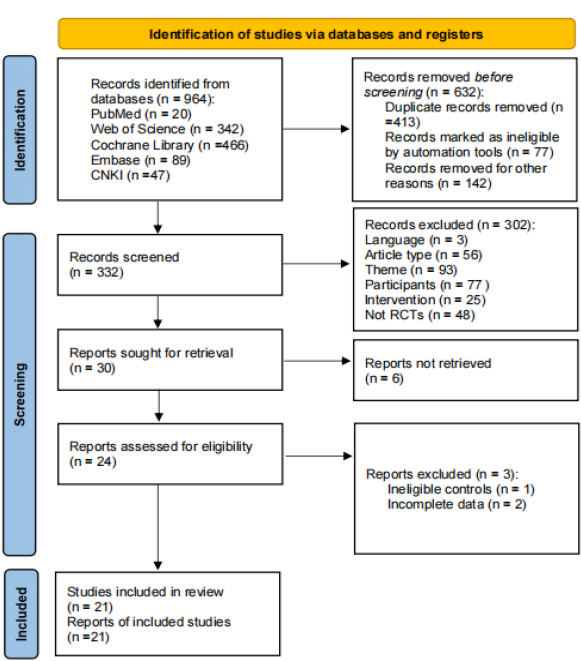
Research and study selection for PRISMA-compliant systematic reviews.

**S4 Table.** Study quality and risk of bias.

| Article | PEDro |  |  |  |  |  |  |  |  |  |  | Sum |
| --- | --- | --- | --- | --- | --- | --- | --- | --- | --- | --- | --- | --- |
|  | 1 | 2 | 3 | 4 | 5 | 6 | 7 | 8 | 9 | 10 | 11 | PEDro |
| Akhtar 2017 | 1 | 1 | 1 | 1 | 1 | 0 | 0 | 1 | 1 | 1 | 1 | 9 |
| ALP 2014 | 1 | 1 | 0 | 1 | 0 | 0 | 1 | 1 | 1 | 1 | 1 | 8 |
| Aluko 2013 | 1 | 1 | 1 | 1 | 0 | 0 | 0 | 1 | 1 | 1 | 1 | 8 |
| Amit 2013 | 1 | 1 | 1 | 1 | 1 | 0 | 0 | 1 | 1 | 1 | 1 | 9 |
| Ferreira 2007 | 1 | 1 | 1 | 1 | 0 | 1 | 0 | 1 | 1 | 1 | 1 | 9 |
| França 2010 | 1 | 1 | 1 | 1 | 0 | 0 | 0 | 1 | 1 | 1 | 1 | 8 |
| Guo 2014 | 1 | 1 | 1 | 1 | 0 | 0 | 0 | 1 | 1 | 1 | 1 | 8 |
| Hlaing 2021 | 1 | 1 | 1 | 1 | 0 | 0 | 0 | 1 | 1 | 1 | 1 | 8 |
| Hosseinifar<br>2007 | 1 | 1 | 0 | 1 | 0 | 0 | 0 | 1 | 1 | 1 | 1 | 7 |
| Inani 2013 | 1 | 1 | 1 | 1 | 0 | 0 | 0 | 1 | 1 | 1 | 1 | 8 |
| Javadian 2012 | 1 | 1 | 0 | 1 | 0 | 0 | 0 | 1 | 1 | 1 | 1 | 8 |
| Macedo 2012 | 1 | 1 | 1 | 1 | 0 | 0 | 0 | 1 | 1 | 1 | 1 | 8 |
| Puntumetakul<br>2013 | 1 | 1 | 1 | 1 | 0 | 0 | 1 | 1 | 1 | 1 | 1 | 9 |
| Shamsi 2016 | 1 | 1 | 0 | 1 | 0 | 0 | 0 | 1 | 1 | 1 | 1 | 7 |
| Sung 2013 | 1 | 1 | 1 | 1 | 0 | 0 | 0 | 1 | 1 | 1 | 1 | 8 |
| Wang 2018 | 1 | 1 | 0 | 1 | 0 | 0 | 0 | 1 | 1 | 1 | 1 | 7 |
| Waseem 2019 | 1 | 1 | 1 | 1 | 0 | 0 | 0 | 1 | 1 | 1 | 1 | 8 |
| Xu 2017 | 1 | 1 | 1 | 1 | 0 | 0 | 0 | 1 | 1 | 1 | 1 | 8 |
| Yang 2022 | 1 | 1 | 1 | 1 | 0 | 0 | 0 | 1 | 1 | 1 | 1 | 8 |
| Zeng 2013 | 1 | 1 | 0 | 1 | 0 | 0 | 0 | 1 | 1 | 1 | 1 | 7 |
| Zhang 2015 | 1 | 1 | 1 | 1 | 0 | 0 | 0 | 1 | 1 | 1 | 1 | 8 |

PEDro-scale-items: 1) Was eligibility criteria specified? 2) Were all subjects randomly allocated? 3) Were allocations concealed? 4) Were the groups similar at baseline? 5) Was there blinding of all participants? 6) Was there blinding of all therapists? 7) Was there blinding of all assessors? 8)Was there measures of at least one key outcome for more than 85% of the subjects initially allocated to groups? 9) Did all subjects for whom outcome measures were available receive the treatment or control condition as allocated or, where this was not the case, data for at least one key outcome was analyzed by "intention to treat"? 10) Were the results of between group statistical comparisons reported for at least one key outcome? 11) Did the study have both point measures and measures of variability for at least one key outcome?

**S5 Figure.**
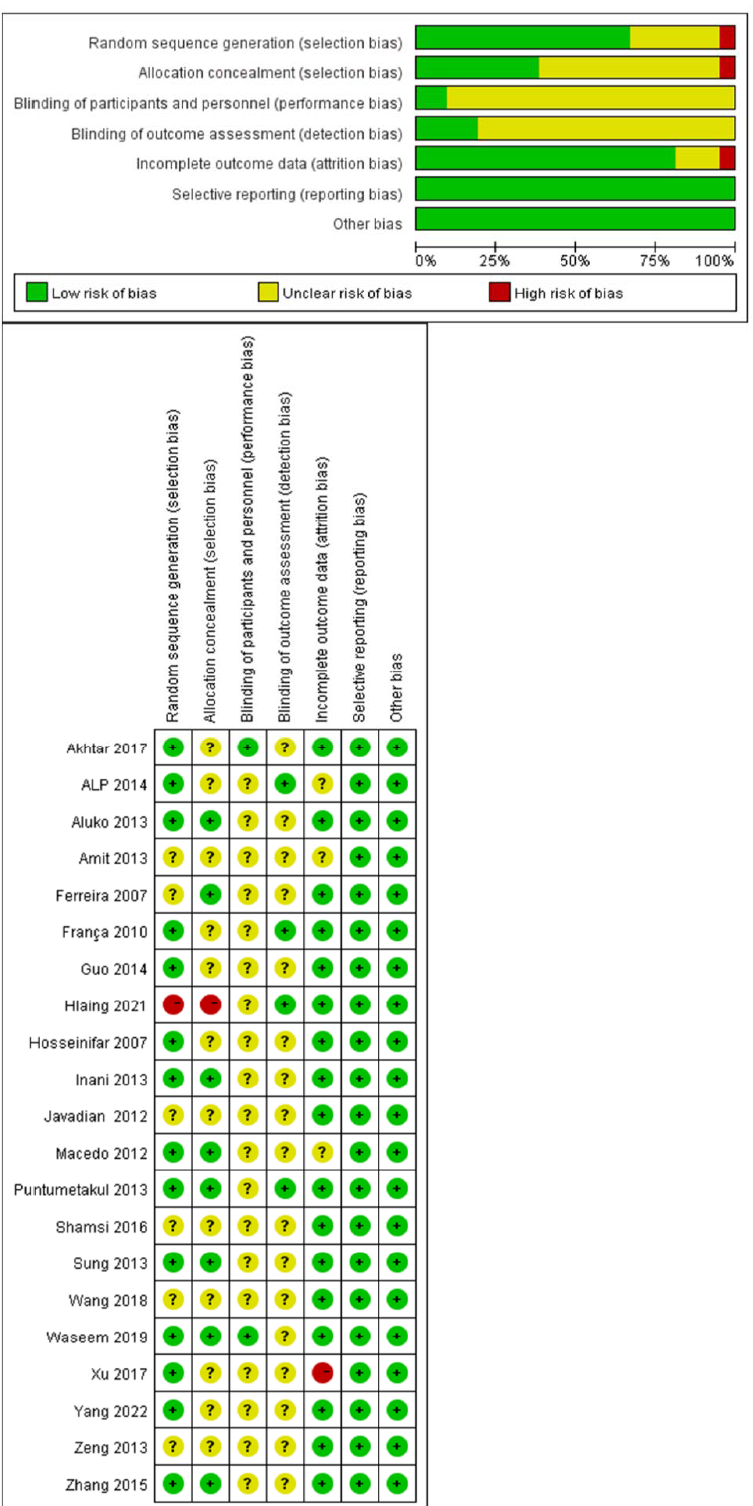
Risk of bias of this review.

**S6 Figure.**
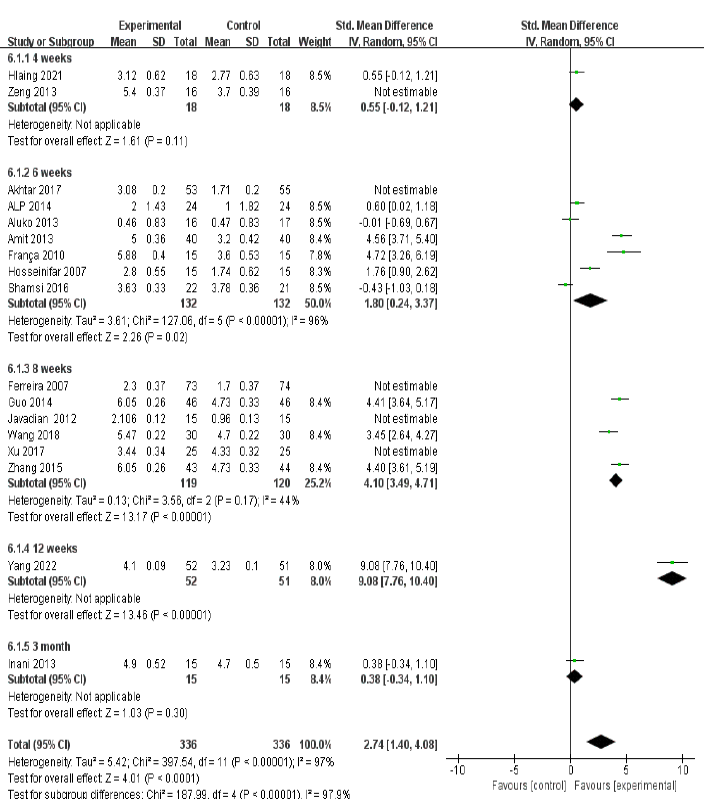
forest plot of pain intensity

**S7 Figure.**
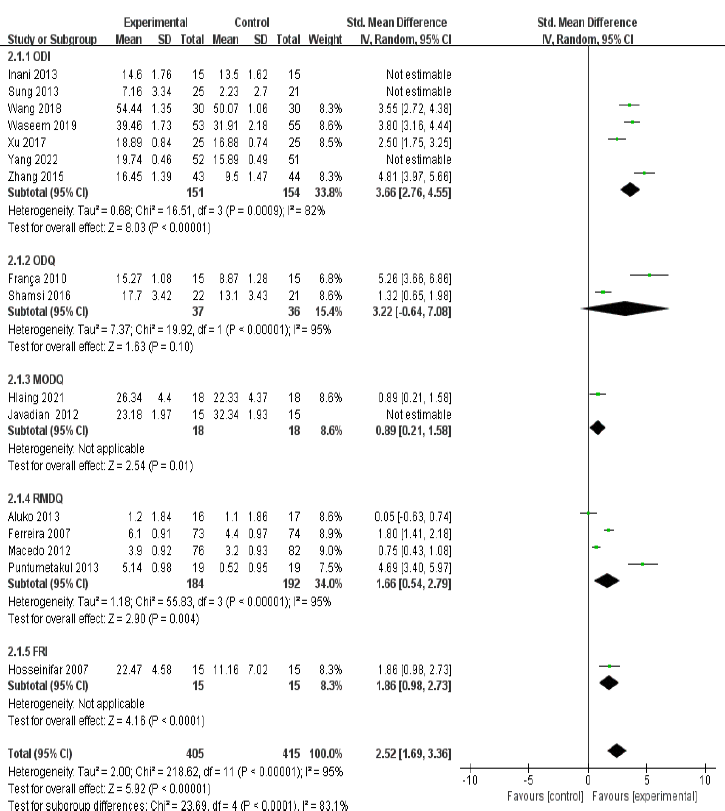
forest plot of disability.

**S8 Figure.**
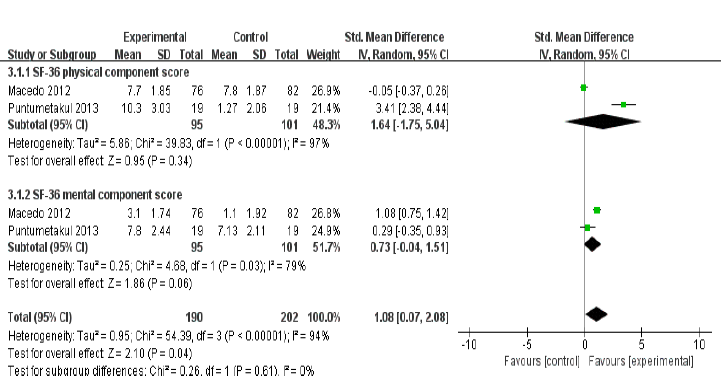
forest plot of generic health.

**S9 Figure.**
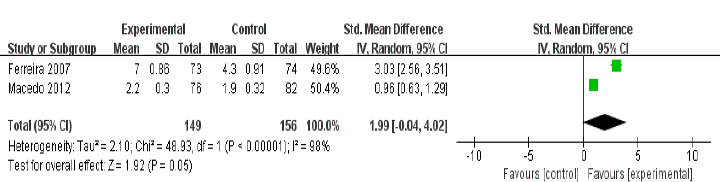
forest plot of specific function.

